# Likelihood of infecting or getting infected with COVID-19 as a function of vaccination status, as investigated with a stochastic model for New Zealand (Aotearoa) for Delta and Omicron variants

**DOI:** 10.1101/2021.11.28.21266967

**Authors:** Leighton M. Watson

## Abstract

**Aim:** The New Zealand government has transitioned from the Alert Level framework, which relied on government action and population level controls, to the COVID-19 Protection Framework, which relies on vaccination rates and allows for greater freedoms (for the vaccinated). Under the COVID-19 Protection Framework and current widespread community transmission of Omicron, there is significant interest in understanding the relative risk of spreading COVID-19 posed by unvaccinated, vaccinated, and boosted individuals.

**Methods:** A stochastic branching process model is used to simulate the spread of COVID-19 for outbreaks seeded by unvaccinated, vaccinated, or boosted individuals. The likelihood of infecting or getting infected with COVID-19 is calculated based on vaccination status. The model is applied to both the Delta and Omicron variants.

**Results:** For the Delta variant a vaccinated traveler infected with COVID-19 is 9x less likely to seed an outbreak than an unvaccinated traveler infected with COVID-19, however, for the Omicron variant there is little difference between outbreaks seeded by unvaccinated and vaccinated individuals (boosted individuals are slightly less likely to seed large outbreaks). For the Delta variant unvaccinated individuals are responsible for 87% of all infections whereas only 3% of infections are from vaccinated to vaccinated when normalized by population. Therefore, a vaccinated individual is 6.8x more likely to be infected by an unvaccinated individual than by a vaccinated individual. For the Omicron variant unvaccinated individuals are responsible for 45% of all infections compared to 39% for vaccinated (two-doses) and 15% for boosted (three-doses) individuals when normalized by population. Despite the vaccine being less effective at preventing breakthrough transmission for Omicron, only 3% of all infections are from boosted to boosted individuals when normalized by population indicating that three doses of the vaccine provide good protection from infection and breakthrough transmission.

**Conclusions:** This work demonstrates that most new infections are caused by unvaccinated individuals, especially for the Delta variant. These simulations illustrate the importance of vaccination in stopping individuals from becoming infected with COVID-19 and in preventing onward transmission. For Omicron, individuals vaccinated with two doses are only slightly less likely to spread COVID-19 than those who are unvaccinated. This work suggests that for the current Omicron outbreak the COVID-19 Protection Framework should potentially be updated to distinguish between those who have received two primary doses of the Pfizer-BioNTech vaccine (vaccinated individuals) and those who have received three doses (boosted individuals).

## Introduction

The 2021 Delta outbreak of COVID-19 in New Zealand caused the government to transition from an elimination strategy to suppression, which relies heavily on vaccination rates. Since the detection of the outbreak on 17 August 2021, double-dose vaccination rates have increased from approximately 22% of the eligible (over 12 years old) population to 93% on 22 January 2022, prior to the detection of Omicron in the community.^1,2,3^ As a result, the COVID-19 pandemic is turning into a pandemic of the unvaccinated; only 11% of hospitalizations in the Delta outbreak were fully vaccinated (defined as more than one week since the second dose of the two-dose Pfizer-BioNTech vaccine).^1,2,3^ Under the COVID-19 Protection Framework, which predominantly uses vaccination certificates, instead of the Alert Level system, which uses population level controls, it is important to understand the relative likelihood of vaccinated/boosted versus unvaccinated individuals spreading COVID-19. This is particularly applicable due to the current widespread community transmission of the Omicron variant.

Here, I use the stochastic model developed in [4] to estimate the likely number of infections caused by an outbreak seeded by an unvaccinated versus vaccinated/boosted individual. This information can help inform reopening decisions and restrictions on travel (e.g., requiring vaccination or a negative test prior to travel). I calculate the likelihood of infecting others or getting infected with COVID-19 based on vaccination status. Mathematical modeling is a useful tool for understanding these probabilities because, as the number of COVID-19 cases in the community has increased, contact tracers have prioritized preventing onwards transmission compared to finding the source of infections.^5^ As a result, the likelihood of infecting others or getting infected as a function of vaccination status is not available for real-world cases but can be determined from model simulations. Results are shown for both the Delta and Omicron variants.

## Methods

A stochastic branching process model is used to simulate the initial spread of a COVID-19 outbreak, similar to previous work by [6-8]. The model tracks the number of infections and the vaccination status of the infecting and infected individuals. The stochastic model used here is the same as presented in [4], which focused on the Delta variant, with the pertinent details summarized below.

### Delta

Each infected individual infects a random number of other individuals, *N*, drawn from a Poisson distribution.^6^ For symptomatic individuals, the Poisson distribution is defined by *λ = RC* where *R* is the reproduction number (chosen to be 6 for the Delta variant) and *C* is the effectiveness of population level controls (e.g., Level 1, 2, 3, or 4 in the Alert Level Framework or Green, Orange, or Red in the COVID-19 Protection Framework).^6^ For an asymptomatic individual, the Poisson distribution is defined by *λ = RC/2*, which assumes that asymptomatic individuals infect, on average, half as many people as symptomatic individuals.^9^ In this work, I only consider *C=1*, which is the situation without any public health measures.

The generation times between an individual becoming infected and infecting *N* other individuals are independently sampled from a Weibull distribution with *a*=5.57 and *b*=4.08 where *a* is the scale parameter and *b* is the shape parameter (mean=5.05 days and variance=1.94 days).^10^ The model assumes that 33% of new infections are asymptomatic (subclinical) with the remainder symptomatic (clinical).^11-13^

The Pfizer-BioNTech vaccine, which is the only COVID-19 vaccine currently being widely administered in New Zealand, is assumed to be 70% effective against infection and 50% effective against transmission for breakthrough infections.^8,14^

High levels of community testing have been essential in identifying cases in the community and, combined with contact tracing and isolation, have been effective at preventing cases that escape from MIQ becoming widespread outbreaks.^15^ It is unclear how testing rates will change in a highly vaccinated public; vaccinated and boosted individuals may feel less need to get tested while unvaccinated individuals may not want to get tested. Therefore, I focus purely on the impact of vaccination rates, particularly on the early stages of an outbreak when cases may be circulating undetected, and following the approach of [4], do not consider testing, contact tracing, or isolation of cases.

Age is not accounted for in this model, either in the vaccination rollout where older individuals are more likely to be vaccinated, or in the susceptibility where older individuals are more likely to experience severe disease or death. Age also plays a role in transmission with young children are less likely to transmit the virus^16,17^ as well as through different ages groups having different levels of mobility and hence different numbers of contacts. See [8, 14] for a New Zealand focused model that accounts for age. Other limitations include not accounting for ethnicity, either in vaccination rates or differential risk factors for different ethnic groups,^18^ or socio-economic status; COVID-19 spreads rapidly through overcrowded households as well as posing a greater risk to those who do not have the economic resources to safely isolate or the ability to work-from-home.^18^

Vaccinated and unvaccinated individuals are modelled as equally likely to interact (based on the vaccination rate). This is a modeling assumption that should be explored further in future work as the COVID-19 Protection Framework and use of vaccine certificates means that in public settings vaccinated individuals are more likely to interact with vaccinated individuals and likewise for unvaccinated individuals. This clustering effect may be more apparent in private gatherings where unvaccinated individuals are potentially more likely to have unvaccinated guests than vaccinated individuals are.

### Omicron

The model described above, originally developed in [4], was focused on the Delta variant. Given the rapid emergence and spread of Omicron in the community, I also apply the model to the Omicron variant with the following modifications.

The population is divided into unvaccinated, vaccinated (two-doses of the Pfizer-BioNTech vaccine), and boosted (three-doses of the Pfizer-BioNTech vaccine). For two doses, the vaccine is modeled as 14% effective against infection of Omicron and 3% effective against transmission for breakthrough infections, which corresponds to 15+ weeks since the second dose was administered. For three doses, the vaccine is 58% effective against infection and 26% effective against breakthrough transmission, which corresponds to 2-5 weeks after the booster was administered. Vaccine effectiveness are taken from [19-21] and are similar to the values used by [22] who modeled an Omicron outbreak in New Zealand. Times are chosen to reflect that many people got their second dose over 3 months ago and that the booster campaign started in earnest a few weeks prior to widespread community transmission of Omicron. Natural immunity from previous COVID-19 infections is not included because, at this stage, the cases numbers are still relatively small compared to the population of New Zealand.

Each infected individual infects a random number of other individuals drawn from a Poisson distribution *λ = R*_*0*_ where *R*_*0*_ is the effective reproduction number in the presence of public health measures (not to be confused with R, which is the reproduction number). Following [22], I use *R*_*0*_ = 2.6, which accounts for the public health measures under the Red level of the COVID-19 Protection Framework (unlike for the Delta variant where I consider R=6 and do not account for public health measures, similar to previous work by [8]). This corresponds to the baseline scenario from [22].

Omicron has a shorter incubation period than Delta^23,24^. For Omicron, the generation time is sampled from a normal distribution with a mean of 3.3 days and standard deviation of 1.3 days.^22^

## Results

### Delta

The simulations are seeded with either one vaccinated (two doses) or one unvaccinated individual at t=0, where t is the time in days, and are run for 31 days (∼1 month) with time steps of 1 day. The simulations are run 100,000 times for each scenario to get a representative sampling of the possible outcomes from the stochastic model.

#### Vaccination Status of Seed Infection

During the Delta outbreak a regional boundary was enforced around Auckland to limit the spread of the COVID-19 outside Auckland, which was the epicenter of the outbreak. The boundary was effective and enabled much of the country to experience minimal restrictions while Auckland was at Alert Level 3 or 4. The regional boundary was relaxed on 15 December 2021 and people were able to travel in and out of Auckland if they were fully vaccinated or had proof of a negative test within 72 hours of traveling.^25^ This situation persisted until 17 January 2022 when the border was removed entirely. Despite these protective measures, the movement of people out of Auckland resulted in COVID-19 being seeded in other locations around the country including Waikato and Northland. Here, I consider the possible numbers of infections in an outbreak based on the vaccination status of the seed infection (Figure 1). The simulations are performed for a vaccination rate of 78.7% of the total population; this is approximately 90% of the eligible population (over 12 years old), which was the government’s vaccination target for all District Health Boards during the Delta outbreak. Note that this work assumes that an infected individual can travel and seed a new outbreak. I do not model the impact of vaccination or testing requirements on preventing infected people from travelling and catching cases before they travel and seed new outbreaks.

**Figure 1:**
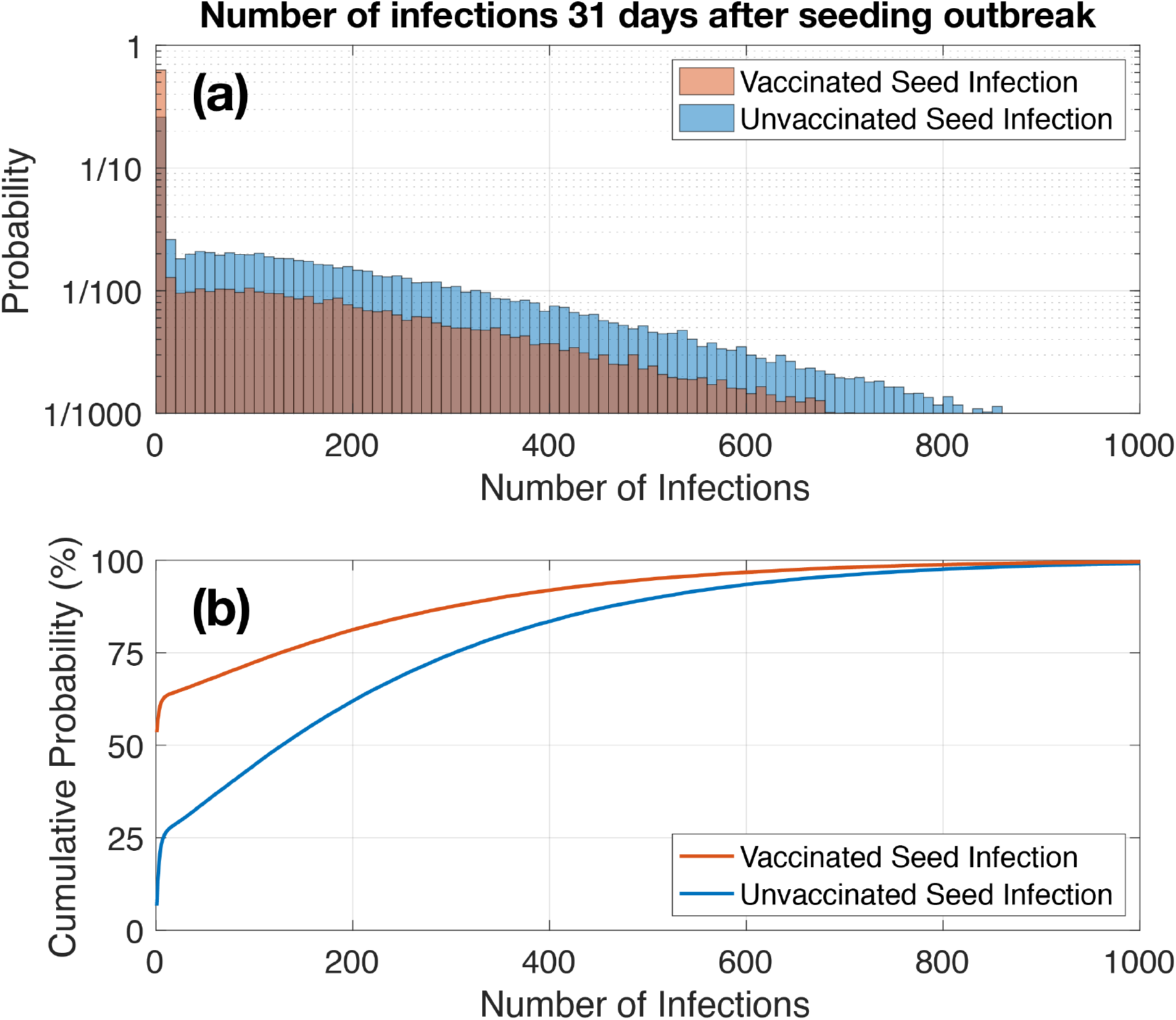
Delta variant. (a) Histograms showing probability of number of infections 31 days into an outbreak and (b) cumulative probabilities for an outbreak seeded by (red) one vaccinated individual or by (blue) one unvaccinated individual.

Figure 1a shows the probability of a given number of infections 31 days into an outbreak seeded by a vaccinated individual or unvaccinated individual, with the cumulative probabilities shown in Figure 1b. For both vaccinated and unvaccinated seed infections, there is a small possibility of large outbreaks developing within the first 31 days of an infection being seeded (5% chance of >506 cases and >657 cases for the vaccinated and unvaccinated seed, respectively). However, for a vaccinated seed infection it is much more likely that COVID-19 does not spread beyond the initial case; for a vaccinated seed infection there is a 54% chance that COVID-19 does not spread to anyone else while for an unvaccinated seed infection there is only a 6% chance. This is because the vaccine is assumed to be 50% effective at preventing onward transmission. ^8,14^ For an unvaccinated seed infection, there is a 54% chance that the outbreak has up to 151 infections after 31 days.

Figure 1 shows the importance of vaccination in stopping outbreaks from being seeded. A vaccinated traveler is 9x less likely to seed an outbreak in a community than an unvaccinated traveler (note that this model does not account for the protection provided by testing requirements prior to traveling, which would reduce the risk factor posed by unvaccinated travelers). This illustrates the importance that travelers are vaccinated (or tested prior to travelling, or both), especially if travelling from regions with significant COVID-19 community transmission (e.g., Auckland) to regions with low vaccination rates (e.g., Northland). Continued community testing (not modeled here) is required to rapidly identify any outbreaks that are seeded before they grow.

#### Likelihood of Infection/Infecting Based on Vaccination Status

The model tracks the number of vaccinated and unvaccinated cases as well as the vaccination status of the individuals that cause the infections. This enables me to calculate the probability of infection based on the vaccination status of the infecting and infected individuals. The results are calculated from the mean of the 100,000 realizations.

Figure 2a shows the results for a vaccination rate of 50% of the total population, which means that there are an equal number of unvaccinated individuals and vaccinated individuals in the population. 87% of all infections are caused by unvaccinated individuals, with 67% of all infections being from unvaccinated individuals to unvaccinated individuals. By contrast, only 13% of infections are caused by vaccinated individuals, and only 3% of these are from vaccinated to vaccinated. This illustrates the importance of vaccination in preventing individuals from (a) getting infected and (b) passing COVID-19 on to others. Figure 2a also illustrates that while the vaccine provides significant protection from getting infected, vaccinated individuals can still get infected, predominantly from unvaccinated individuals. Vaccinated individuals are 6.8x more likely to be infected by an unvaccinated individual than by a vaccinated individual. Although Figure 2a is calculated for a vaccination rate of 50%, it also gives the values that would be observed at different vaccination rates after normalizing for population (dividing the number of infections by the number of people in each category). Normalizing by population removes the influence of the number of people in each category (for example, more cases amongst vaccinated individuals than unvaccinated, which occurs simply because there are many more vaccinated people). This illustrates that most infections are caused by unvaccinated individuals.

**Figure 2:**
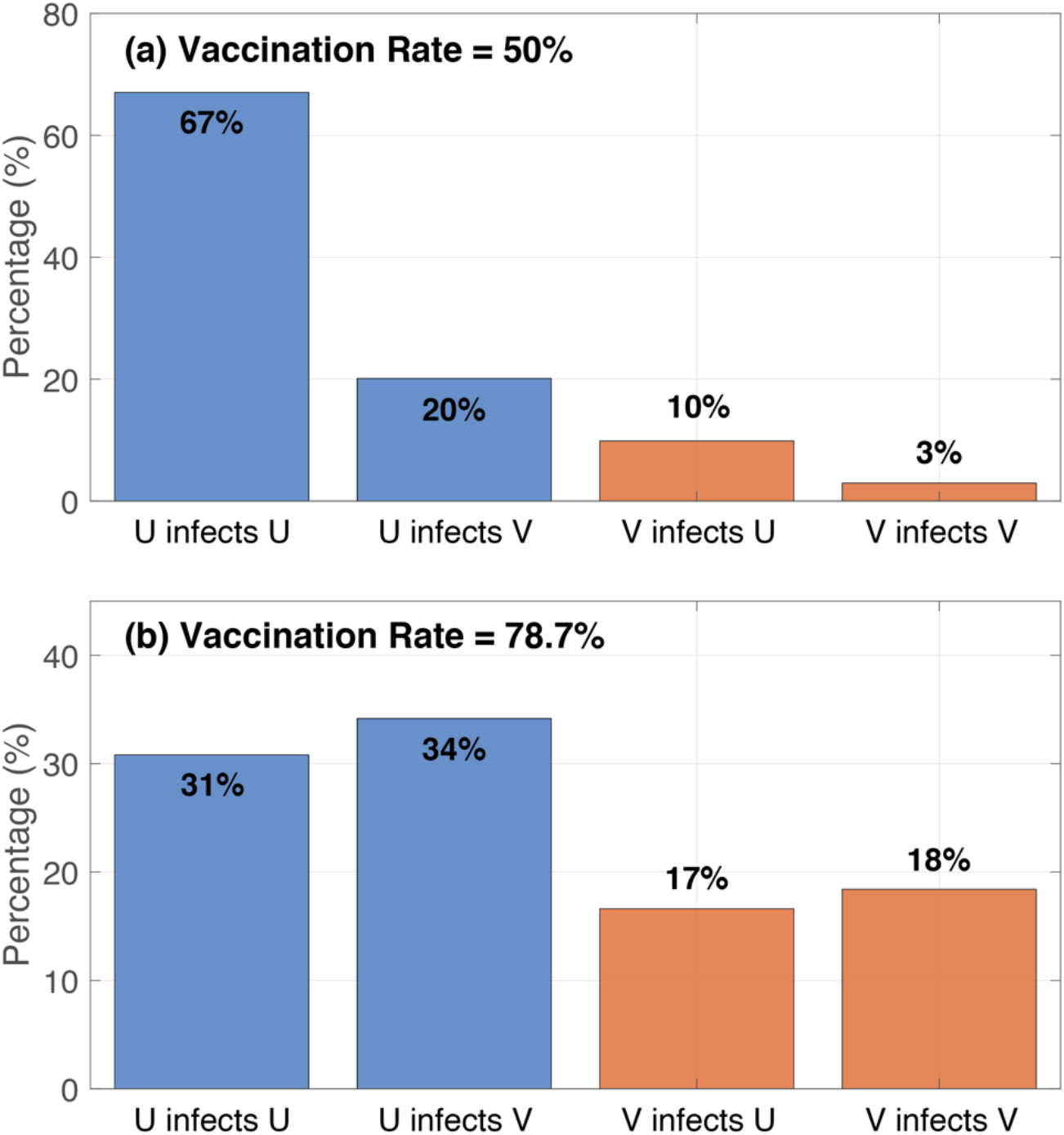
Delta variant. Likelihood that a new infection is caused by a (U) unvaccinated or (V) vaccinated individual and that the new infection is in an unvaccinated or vaccinated individual. (a) shows a total vaccination rate of 50%, where there are an equal number of vaccinated and unvaccinated individuals in the population, while (b) shows a total vaccination rate of 78.7%, which is approximately the 90% eligible population target. (a) shows the expected result when normalizing by population.

Figure 2b shows the results for a total vaccination rate of 78.7%. Unvaccinated individuals are responsible for 65% of infections despite only making up 21.3% of the population. By contrast, vaccinated individuals are only responsible for 35% of infections while making up 78.7% of the population. Even at these high levels of vaccination where there are 3.7x as many vaccinated individuals are unvaccinated individuals, a new infection is almost twice as likely to be caused by an unvaccinated individual. A vaccinated individual has a 65% chance of being infected by an unvaccinated individual compared to a 35% chance of being infected by a vaccinated individual (1.9x more likely to be infected by unvaccinated individual even though there are far fewer unvaccinated individuals in the population). Figure 2b illustrates that, even at high levels of vaccination, unvaccinated individuals are the main cause for continued spread of COVID-19 with only 18% of infections from vaccinated to vaccinated. This suggests that restricting unvaccinated individuals from high-risk locations (i.e., potential super-spreader events) will help to minimize the spread of COVID-19, which is the goal of the COVID-19 Protection Framework.

The model assumes that unvaccinated and vaccinated individuals are equally likely to interact. In reality, unvaccinated and vaccinated individuals are likely to interact with individuals with the same vaccination status, both in private gatherings and in public spaces as mandated by the COVID-19 Protection Framework. Therefore, the results in Figure 2 may underestimate the spread of COVID-19 between unvaccinated individuals.

### Omicron

The simulations are seeded with either one boosted (three doses), vaccinated (two doses), or unvaccinated individual at t=0, where t is the time in days, and are run for 31 days (∼1 month) with time steps of 1 day. The simulations are run 100,000 times for each scenario to get a representative sampling of the possible outcomes from the stochastic model. I consider a population that is 10% unvaccinated, 40% vaccinated (two doses) and 50% boosted (three doses), although the booster uptake is varied in Figure 4 to examine the effect of increasing booster coverage.

#### Vaccination Status of Seed Infection

I perform a similar analysis to Figure 1 for the Omicron variant and consider outbreaks seeded by unvaccinated, vaccinated, and boosted individuals with the results shown in Figure 3. Figure 3a shows the probability of a given number of infections 31 days into an outbreak as a function of the vaccination status of the seed infection while Figure 3b shows the cumulative probabilities. The model assumes that the Pfizer-BioNTech vaccine is significantly worse at preventing infection and transmission against Omicron than Delta (two-dose are assumed to be 70% effective against infection and 50% effective against transmission for Delta but only 14% and 3%, respectively, against Omicron). Therefore, there is not much difference between the unvaccinated and vaccinated seed infections (69% and 71% chance of less than 500 infections after 31 days for unvaccinated and vaccinated seeds, respectively). Three doses of the Pfizer-BioNTech vaccine provide decent protection from infection and transmission (58% and 26%, respectively), although three doses are still less effective against Omicron than two doses were against Delta. Note that vaccine effectiveness wanes with time and that model results shown here are only applicable for a snapshot in time.

**Figure 3:**
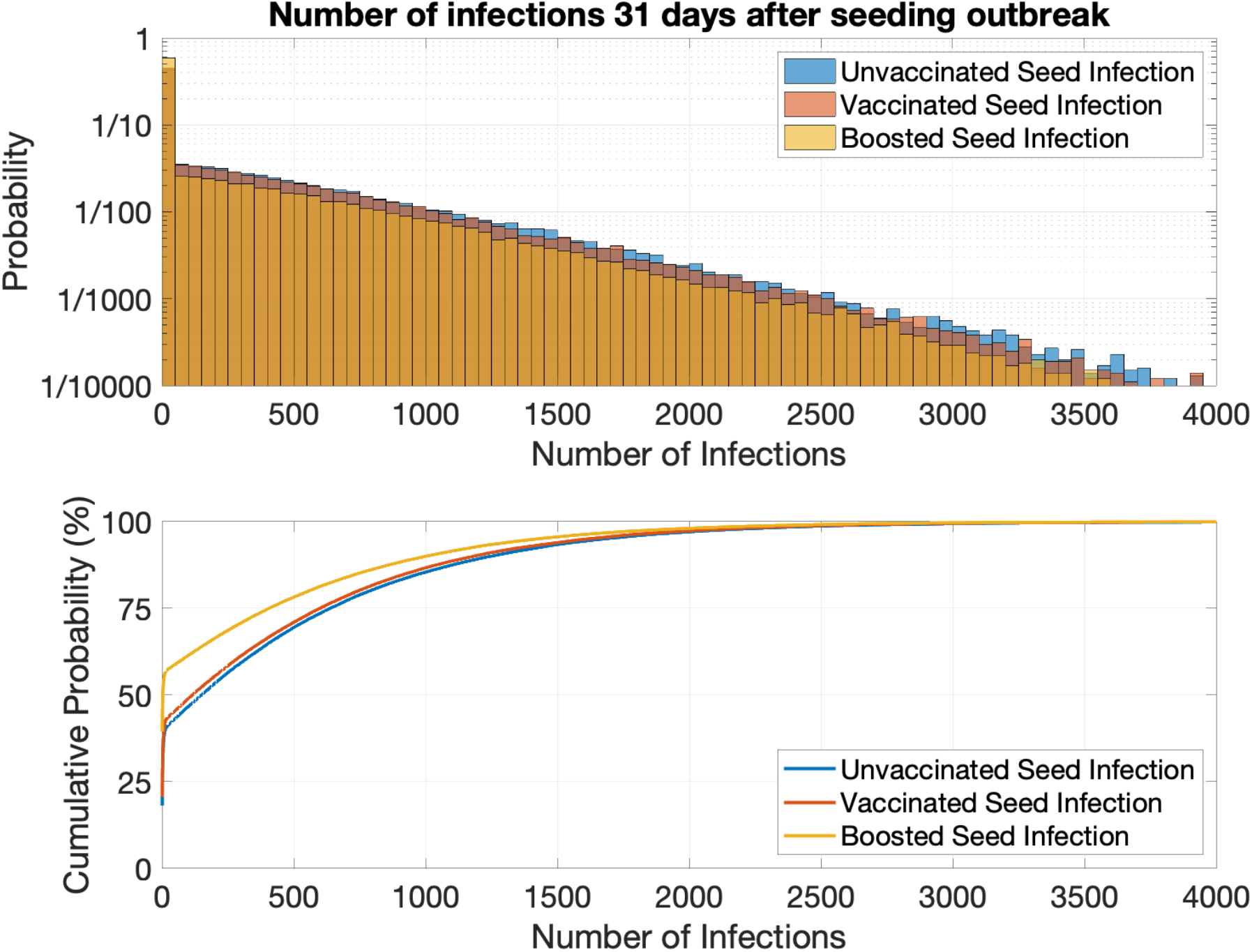
Omicron variant. (a) Histograms showing probability of number of infections 31 days into an outbreak and (b) cumulative probabilities for an outbreak seeded by (blue) one unvaccinated, (red) one vaccinated, and (yellow) one boosted individual.

For a boosted seed infection, there is a 78% chance of less than 500 infections are 31 days. Boosted seed infections are more likely to lead to small outbreaks that self-extinguish after a small number of infections. An outbreak that starts from a boosted individual has a 56% chance of stopping after 10 or fewer infections whereas the probability is only 42% for a vaccinated seed infection or 40% for an unvaccinated seed infection.

This demonstrates that while the Pfizer-BioNTech vaccine is less effective against preventing infection and transmission of Omicron compared to Delta, those who are boosted are slightly less likely to seed a large outbreak.

#### Booster Vaccination Rate

I also consider the impact of increasing booster rates from 50% to 70% and 90% (assuming 10% of the population remain unvaccinated). The results are shown in Figure 4 for an unvaccinated seed infection. Increasing the percentage of the population who are boosted drastically reduces the likely number of infections. For a booster rate of 50% there is an 47% chance of less than 100 cases after 31 days compared to 77% and 98% for booster rates of 70% and 90% respectively. This clearly illustrates that while three doses of the vaccine provide imperfect protection (the model presented here assumes 56% effective against infection and only 26% effective against breakthrough transmission), high levels of booster coverage provide good protection against infection on a population level. This demonstrates the need for everyone to get boosted to provide the best possible protection for themselves and their community.

**Figure 4:**
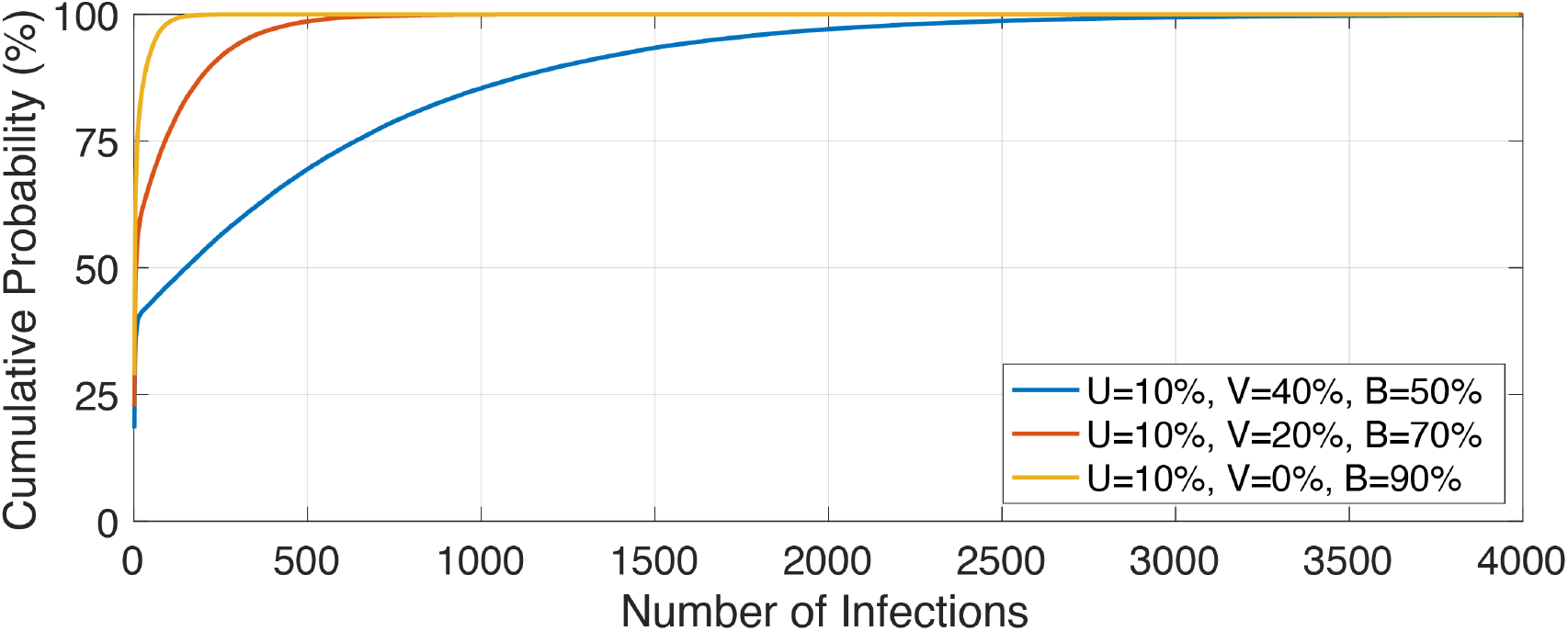
Cumulative probability for number of infections 31 days after seeding an outbreak with an unvaccinated seed infection for booster vaccination rate of (blue) 50%, (red) 70%, and (yellow) 90%.

#### Likelihood of Infection/Infecting Based on Vaccination Status

The Omicron version of the model tracks the number of unvaccinated, vaccinated, and boosted cases as well as the vaccination status of the individuals that cause the infections. Like the results shown in Figure 2 for the Delta variant, I calculate the probability of infection based on the vaccination status of the infecting and infected individuals. The results are shown in Figure 5 and are calculated from the mean of the 100,000 realizations for an outbreak seeded with one unvaccinated individual, one vaccinated, and one boosted.

**Figure 5:**
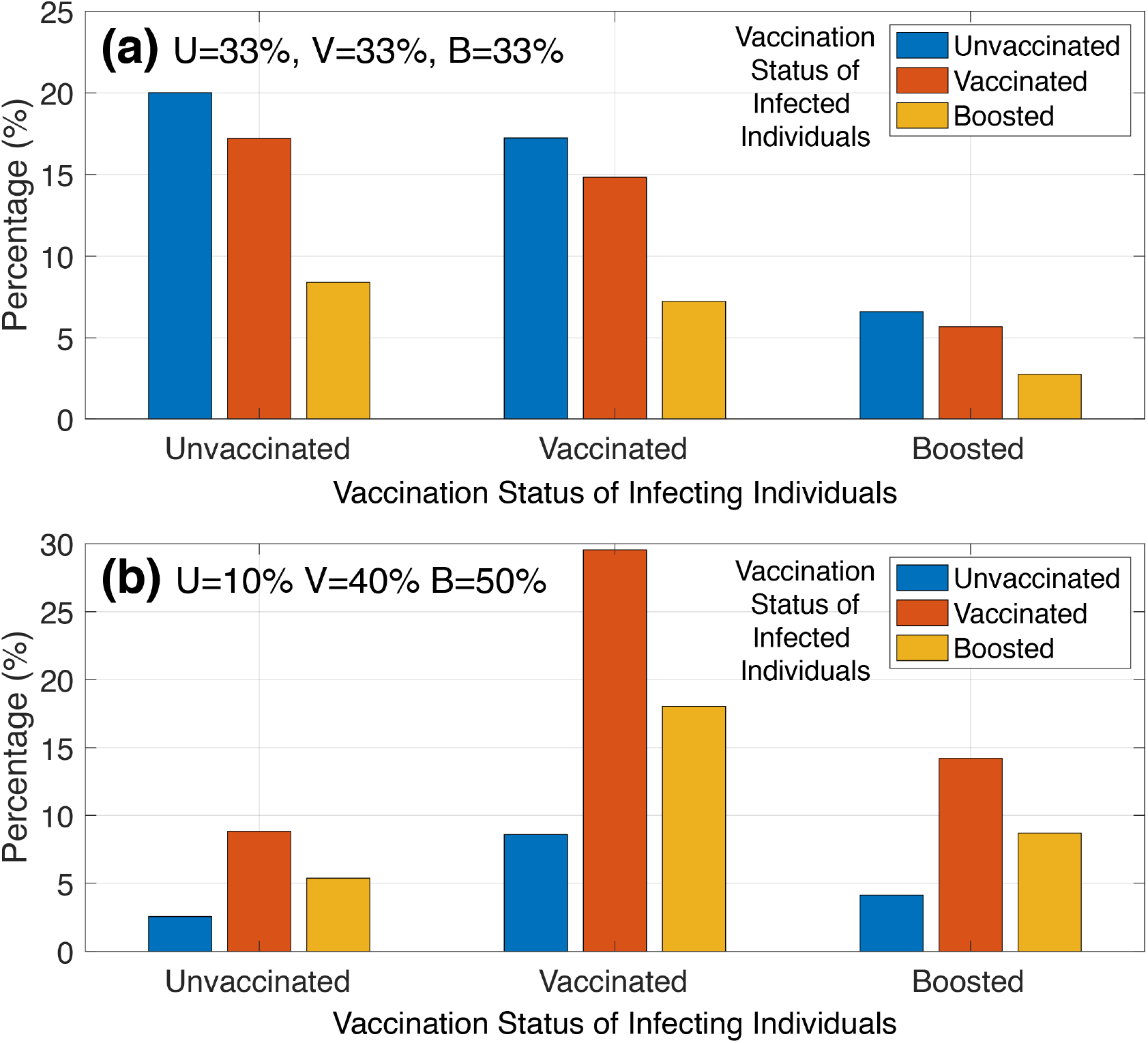
**Omicron** variant. Likelihood of new infections as a function of vaccination status of the infecting and infected individuals. (a) Vaccination rates of U=33% (unvaccinated), V=33% (vaccinated), and B=33% (boosted). This shows the expected result when normalizing by population. (b) Vaccination rates of U=10% (unvaccinated), V=40% (vaccinated), and B=50% (boosted).

Figure 5a shows the result for when there are an equal number of unvaccinated, vaccinated, and boosted individuals in the population. This is also the result expected when normalized for population (dividing the number of infections by the number of people in each category). Unvaccinated individuals are responsible for 46% of infections compared to 39% for vaccinated and 15% for boosted. Of the new infections, unvaccinated individuals make up 44% of new cases while vaccinated are 38% and boosted are 18%. Unvaccinated individuals are 3.1x more likely to infected others and 2.4x more likely to be infected than boosted individuals. This supports the rationale of using the COVID-19 Protection Framework to restrict unvaccinated individuals from high-risk settings, although the definition of “fully vaccinated” should be updated to distinguish between individuals who have had two or three primary doses of the vaccine. When normalized by population (as shown in Figure 5a), transmission from boosted to boosted individuals is only responsible for 3% of infections. These results are sensitive to the model assumptions about vaccine effectiveness but nonetheless shows that even through the Pfizer-BioNTech vaccine is less effective against Omicron than Delta (particularly for two doses), boosted individuals are much less likely to spread COVID-19 or be infected with COVID-19. Vaccinated individuals are only slightly less likely to spread COVID-19 or get infected when compared to unvaccinated, demonstrating the need to get boosted to protect against Omicron infections and prevent onward transmission.

Figure 5b shows the result for realistic vaccination rates of 10% unvaccinated, 40% vaccinated, and 50% boosted. In this situation unvaccinated individuals are responsible for 17% of infections compared to boosted individuals who are responsible for 27% of infections. 15% of new infections occur in unvaccinated individuals compared to 32% in boosted individuals. The boosted population is 5x larger than the number of unvaccinated individuals but only responsible for causing 1.6x as many infections and receiving 2.1x as many (much less than the 5x as many that would be expected if the vaccine did not offer any protection). This illustrates that three doses of the Pfizer-BioNTech vaccine are effective at preventing infection and transmission.

## Conclusions

The New Zealand government has transitioned from the Alert Level system to the COVID-19 Protection Framework that replaces population level controls with vaccination certificates. As a result, there is a need to better understand the risk posed by unvaccinated versus vaccinated and boosted individuals. Here, I use a stochastic model to simulate the potential number of infections in an outbreak seeded by a unvaccinated individual versus a vaccinated individual (for Delta) and a boosted individual (for Omicron). For Delta, unvaccinated individuals are much more likely to seed an outbreak with a 54% chance of causing an outbreak with over 107 cases after 31 days. By contrast, for a vaccinated seed infection, there is a 54% chance that the outbreak does not spread beyond the initial seed. Vaccinated travelers are 9x less likely to seed an infection than unvaccinated travelers. For Omicron, there is little difference between unvaccinated and vaccinated seed infections but boosted individuals are slightly more likely to result in small outbreaks that self-extinguish. Increasing booster coverage can significantly slow the growth of an outbreak and makes it much more likely that outbreaks will self-extinguish after a small number of cases.

I also calculate the likelihood of getting infected and of infecting others based on vaccination status for Delta and Omicron variants. For Delta, unvaccinated individuals are much more likely to spread the virus and, when normalized by population, are responsible for 87% of all. Transmission between vaccinated individuals is rare and responsible for only 3% of all infections when normalized by population. The Pfizoer-BioNTech vaccine is less effective against Omicron. Nonetheless, unvaccinated individuals are more likely to spread the virus and are responsible for 46% of new infections compared to 39% for vaccinated individuals (two-doses) and 15% for boosted individuals (three-doses), when normalized by population. Despite three-doses of the vaccine having limited effectiveness against breakthrough transmission (26%), transmission between boosted individuals is rare accounting for only 3% of all infections when normalized by population. This illustrates that COVID-19 is becoming a pandemic of the unvaccinated and is predominantly spread by the unvaccinated, especially for the Delta variant. For Omicron, the two-dose vaccinated individuals are only slightly less likely to be infected and infect others than the unvaccinated but those who are boosted (three doses) are much less likely to be infected or infect others.

The COVID-19 Protection Framework and associated vaccine certificates distinguish between unvaccinated and vaccinated individuals. This work suggests that for the current Omicron outbreak, the COVID-19 Protection Framework should potentially be updated to further distinguish between those who have received two primary doses of the Pfizer-BioNTech vaccine (vaccinated individuals) and those who have received three doses (boosted individuals).

## Data Availability

The MATLAB code produced in the present study is available upon reasonable request to the author.

